# Geographical identification of the vulnerable groups during COVID-19 crisis: the typhoon eye effect and its boundary conditions

**DOI:** 10.1101/2020.04.28.20083667

**Authors:** Pok Man Tang, Stephen X. Zhang, Chi Hon Li, Feng Wei

## Abstract

**Aim:** Although some studies suggest the coronavirus disease (COVID-19) is associated with negative consequences on physical health, our knowledge about the detrimental effects of COVID-19 on people’s mental health is still nascent. This study uses typhoon eye theory to offer insights in helping clinical psychiatrists to screen people with well-being issues during COVID-19 outbreak.

**Methods:** We collected survey data from working adults across different geographical areas in China on 20 and 21 February 2020 during the outbreak of COVID-19. The sample contains 308 working adults, who were in various parts of China, with varying distance to the epicenter of Wuhan.

**Results:** Individual adults’ distance to the epicenter was negatively associated with life satisfaction (β = −0.235, 95% CI −0.450 to −0.020, *p* = 0.032). This association between distance and life satisfaction was significant only for adults who were young or had smaller family sizes. For example, the negative relationship was strongest when the individuals were in the age bracket of 20 years old (15.7%; β = −0.703, 95% CI −1.098 to −0.307; *p* = 0.001) and single (32.3%; β = −0.767, 95% CI −1.125 to −0.408; *p* < 0.001).

**Conclusion:** Our results that people’s well-being deteriorates by the distance from the epicenter for specific groups of people help guide mental healthcare providers towards the regions that are further away from the epicenter in the ongoing COVID-19 outbreak. Meanwhile, our results indicate the practitioners should be cautious of using typhoon eye effect for individuals who were older or had a larger family size.

## INTRODUCTION

The COVID-19 virus is deteriorating people’s wellbeing, as shown in an increase in the burnout or anxiety across communities.^1 2^ However, our knowledge of people’s life satisfaction, one of the most prominent indicators of mental health, ^3 4^ during COVID-19 crisis remains a lacuna. We aim to use early evidence in China to help mental health services providers in screening people with wellbeing issues during COVID-19 outbreak from a novel perspective of typhoon eye theory.^5 6^ This study identifies the vulnerable regions where individuals are more likely to suffer from well-being issues and helps to guide medical professionals’ attention towards the more mentally vulnerable groups that are far away from the epicenter in the COVID-19 outbreak.

Mental health services used to pay more attention to people around the center of the epicenter following the “ripple effect” during severe acute respiratory syndrome (SARS) and Ebola ^7–9^ However, given the explosive information on COVID-19,^10^ we suspect that individuals’ well-being deteriorates over distance from the epicenter, as depicted by the psychological typhoon eye theory.^5 6^ To provide evidence on the geographical distribution of the wellbeing issues during COVID-19 crisis, we test whether typhoon eye theory works and for whom it works in the COVID-19 outbreak.

To further help clinical psychiatrists in conducting clinical screening for the vulnerable groups of people during the pandemic of COVID-19, we examine whether the typhoon eye effect is more or less useful based on people’s age and family size. The younger population are usually more adaptive to the outbreak of virus or a natural disaster,^11 12^ however, they also tend to access information on COVID-19 more frequently via social media and other digital channels^13^ and hence are more exposed to the associated negative content.^14^ In addition, family size is a salient indicator of social support that one could receive.^15^ During the outbreak of a disease, family size has been found to become an important resource to buffer stress and anxiety.^16^

We surveyed working adults in locations that vary in their travel distance from the epicenter of Wuhan (0 to 2126 km) to examine its relationship with their life satisfaction. Moreover, we have also identified the vulnerable groups of people (based on age and family size), for whom the typhoon eye effect of COVID-19 is more pronounced. Overall, drawing from psychological typhoon eye,^5 6^ this study provided a snapshot of adults’ well-being during the ongoing COVID-19 pandemic to enable more targeted mental health support.

## METHODS

### Study design

On December 29, 2019, the first four cases of COVID-19 were identified by the local hospitals in Wuhan, Hubei Province, China.^17^ Given the severity of COVID-19, the World Health Organization made the decision on presenting the outbreak as a Public Health Emergency of International Concern (PHEIC) on January 30, 2020.^18^ A week prior to this announcement, the Chinese authorities decided to shut down the city of Wuhan by ceasing all the transportation means, including flights, trains, subway, and ferry services.^19^ Nevertheless, around 300,000 people traveled out of Wuhan area by train before the policy was implemented.^20^ About 70% of those people reached reach other areas within Hubei Province, and about 14% went to the nearby provinces, such as Henan, Hunan, Anhui, and Jiangxi.^21^ As people left the epicenter, it is susceptible that they carry fear and anxiety to people around them due to the concerns about being infected by the virus.

### Data collection and measures

From February 20 to 21, 2020, we sent a survey to 410 working adults staying in various cities in China right after a month of the outbreak. 308 of them answered the survey, resulting in a response rate of 75.1%. Their distance to the epicenter ranged from 0 km to 2126 km. All respondents agreed to participate in the study, which was approved by the ethics committee at Tongji University (#20200211).

Our survey gathered subjects’ life satisfaction, distance to the epicenter, and socio-demographic characteristics, including gender, age, education, family size, the number of office days last week, and job status. Life satisfaction was assessed with the Satisfaction With Life Scale (SWLS),^22^ which has five items (Cronbach’s alpha = 0.72) shown in Table 1. The distance to the epicenter was calculated as each participant’s distance to Wuhan at the time of the survey. Education included the categories of “elementary school”, “middle school”, “high school”, “vocational school”, “bachelor”, “master”, and “doctorate”. Family size was measured as “single”, “married (without a child)”, “married (with 1 child)”, “married (with more than 1 child)”, and “others: divorced/widowed”. Job status was classified as “usual work routine”, “home office”, and “work suspended”.

**Table 1.** Scale of life satisfaction

| Items |
| --- |
| 1. In most ways my life is close to my ideal. |
| 2. The conditions of my life are excellent. |
| 3. I am satisfied with my life. |
| 4. So far I have gotten the important things I want in life. |
| 5. If I could live my life over, I would change almost nothing. |

We have also conducted a supplementary analysis on the relationship between the distance to the epicenter and life satisfaction from data collected prior to the outbreak of COVID-19 to check whether the typhoon eye effect occurred only during the COVID-19 crisis.

We used multiple linear regression to predict life satisfaction as an outcome by subjects’ gender, age, family size, education, number of working days last week, job status, and distance to the epicenter. We reported the descriptive and regression findings on unweighted data with STATA 16.0 by the statistical significance of p < 0.05 in Table 2.

**Table 2.**
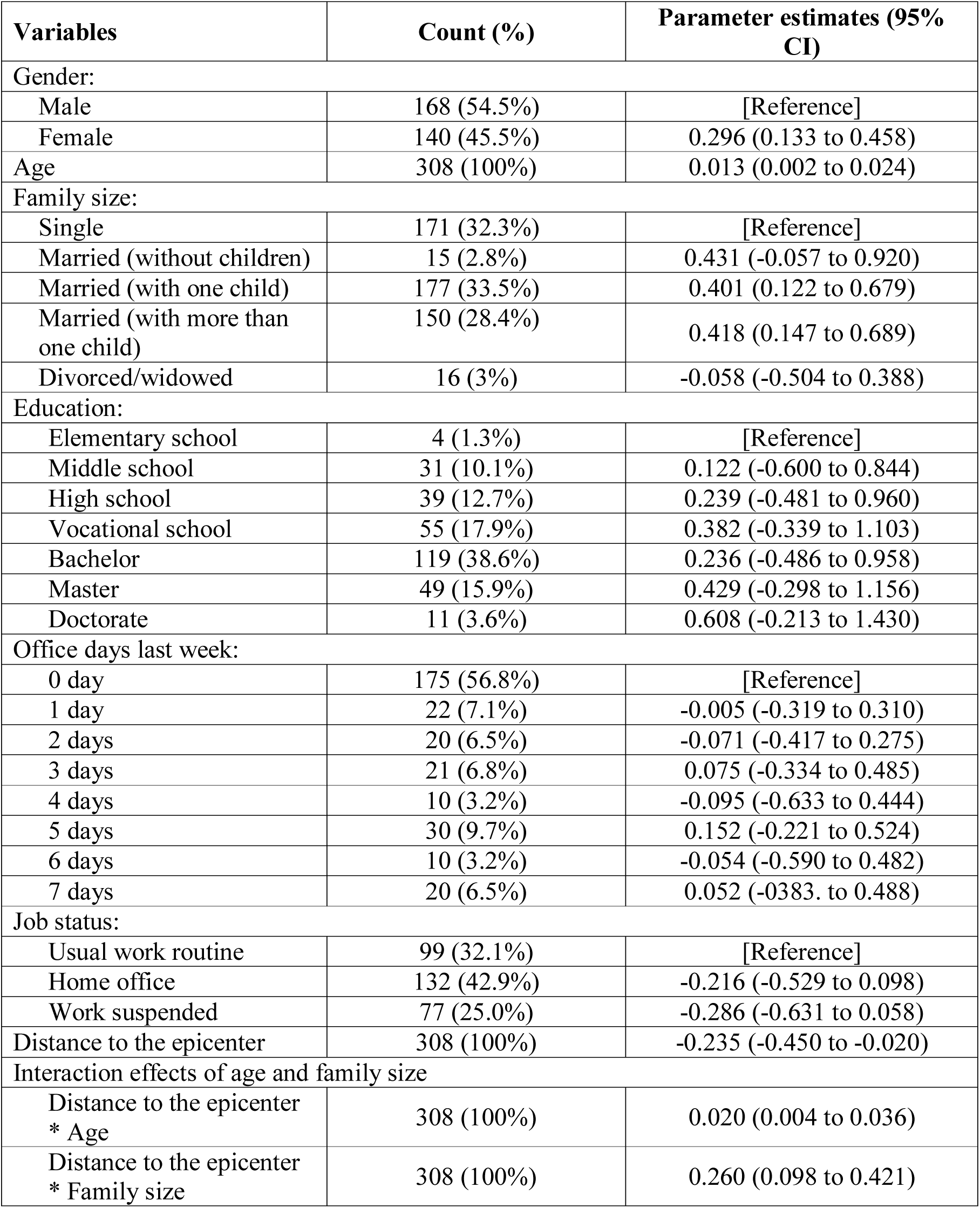
Mean estimated life satisfaction among subjects based on their gender, age, education, family status, marriage status, and the distance to the epicenter

## RESULTS

Around half 168 (54.5%) of the subjects were male, and 140 (45.5%) were female. The average distance to the epicenter of Wuhan was 0.81 (in 1000 km) with a standard deviation of 0.41 (in 1000 km). The average age of the subjects was 38 (s.d.= 9). For family size, most of them fell into the categories of single: 171 (32.3%), married with one child: 177 (33.5%), and married with more than one child: 150 (28.4%). There were 119 (38.6%) people with bachelor’s degrees and 129 (42%) with elementary, middle, high, and vocational school education levels. During our survey, more than half of them (175, 56.8%) did not have any office day in the past week and the rest worked on average for 1.67 (s.d. 2.34) days last week. 22 (7.1%) worked one day and 20 (6.5%) worked seven days in the past week. 132 (42.9%) worked from home, while 99 (32.1%) maintained the usual work routine, and 77 (25%) suspended their work.

### Main Effects of the Personal Characteristics

The regression results in Table 2 examined the predictors of life satisfaction during the period of the outbreak. Male (54.5%) reported a lower level of life satisfaction than women did (45.5%) (β = 0.3, 95% CI 0.13 to 0.46, *p* < 0.001). Younger working adults experienced less life satisfaction than older adults did (β = 0.013, 95% CI 0.002 to 0.024; *p =* 0.019). Single people had less life satisfaction compared to those with one child (33.5%) and more than one child (28.4%) (for one child: β = 0.401, 95% CI 0.122 to 0.679; *p <* 0.005; for more than one children: β = 0.418, 95% CI 0.147 to 0.689; *p =* 0.003). The effects of education, the number of office days last week, and job status on life satisfaction were not significant.

### Effect of the Distance to the Epicenter of Wuhan

The regression results in Table 2 indicated the negative association between the distance to the epicenter and life satisfaction (β = −0.235, 95% CI −0.450 to −0.020; *p =* 0.032), supporting the typhoon eye effect.

First, the association between the distance to the epicenter and life satisfaction was less negative among the older adults (β = 0.020, 95% CI 0.004 to 0.036; *p =* 0.015). The association is the most negative in the youngest age bracket of 20 years old (β = −0.703; 95% CI −1.098 to −0.307; *p =* 0.001) and the second most negative in the second youngest age bracket of 30 years old (β = −0.475, 95% CI −0.748 to −0.202, *p =* 0.001), while the group of 40 years old were the third one (β = −0.247, 95% CI −0.460 to −0.034; *p =* 0.023). The association was not significant for the age brackets of 50 years old and above. These interaction effects are shown in Figure 1.

**Figure 1.** The association between distance to the epicenter and life satisfaction by individuals’ age. The interaction effect was significant and strongest in the age brackets of 20, followed by the brackets of 30 and 40. The interaction effect was insignificant in the brackets of above 50.

Second, the negative association between the distance to the epicenter and life satisfaction was also buffered by large family size (β = 0.260, 95% CI 0.098 to 0.421; *p =* 0.002). Subgroup analysis revealed the typhoon eye effect was significant among the singles (β = −0.767, 95% CI −1.125 to −0.408; *p <* 0.001), those married without a child (2.8%; β = - 0.507, 95% CI -.0750 to −0.265; *p <* 0.001), and those married with one child (33.5%; β = −0.248, 95% CI −0.451 to −0.044; *p =* 0.017). The association between the distance to the epicenter and life satisfaction was not significant for those married with more than one child (25.4%; β = 0.012, 95% CI −0.264 to 0.288; *p =* 0.932) or indicated others in family status (3%; β = 0.272, 95% CI −0.132 to 0.675; *p =* 0.186). These interaction effects are demonstrated in Figure 2.

**Figure 2.** The association between distance to the epicenter and life satisfaction by individuals’ family size. The interaction effect was significant and strongest for the individuals who were single, followed by those who married without a child and with a child. The interaction effect was insignificant for those who married with more than one child and others.

### Supplementary Analysis

We modeled the relationship between the distance to the epicenter and life satisfaction using collected data from working adults before the outbreak of COVID-19. The effect of the distance to the epicenter of Wuhan on life satisfaction was insignificant (β = 0.000, 95% CI - 0.0003 to −0.0003, p = 0.897) before the outbreak. This supplementary analysis indicates that the typhoon eye effect did not occur before the COVID-19 outbreak.

## DISCUSSION

This study shows that people who were geographically further from the epicenter experience lower levels of life satisfaction in China. Following psychological typhoon eye theory, we further identify the vulnerable groups that mentally suffer the most associated with their distance during the outbreak: *younger adults* and individuals with *small family size* to provide important insights for mental health service providers while conducting the clinical screening. In summary, our findings do not only provide psychiatric implications on the location of individuals from the outbreak area but also combine this geographical result with the demographic characteristics of participants to help identify the vulnerable groups who were in greater need of medical attention.^23^

Indeed, our results empirically confirmed the postulation of the typhoon eye theory.^5 6^ Research from multiple disciplines of research can provide explanations of the findings. For example, clinical studies found that people that did not experience the situation first-hand often overestimated the likelihood of infections.^24^ People who learned the news about the virus away from the epicenter tended to exacerbate their concerns over COVID-19.^25^ Similarly, research after the 2008 Wenchuan earthquake in China found that people far from the earthquake area were more likely to suffer mentally for their higher estimation of post-earthquake concern.^5^ Understanding the life satisfaction of people with a varying geographical distance to the epicenter is crucial for clinical psychiatrists during the process of screening because the changes in its score are associated with the chance of suffering from mental disorders like depression.^26 27^ By revealing that people who were more distant from the epicenter indeed have lower life satisfaction scores, our results guide mental health professionals to prioritize the treatment and habilitation for people based on their geographical distance to the outbreak area.

The findings that the typhoon eye effect was more pronounced among younger are consistent with prior research. As noted earlier, younger people tend to rely on social media and the internet to obtain information.^28^ This reliance increases the likelihood of developing the feelings of anxiety and burnout through inauthentic news about the epicenter. Younger residents can be more vulnerable to negative information on COVID-19. The other finding that the psychological typhoon eye effect is more salient for people with smaller family sizes consonants with the family literature.^29^ Support from the family attenuates the negative psychological impacts that one might experience facing disasters.^30^ Hence, we push forward the claim that one’s family size is a proxy of the level of social and emotional support that individuals may receive during the outbreak of COVID-19 ^31^.

Practically, our findings combine geographical and demographic information of participants to help identify the vulnerable groups who were in a greater need for mental health attention. Our results suggest that mental health services cannot solely use typhoon eye effect as the only geographical information to identify those with low life satisfaction. The results that only people in the age brackets of 20–40 in regions far from the epicenter were more likely to experience lower levels of life satisfaction indicate that mental healthcare providers can indeed use typhoon eye effect only for people in those age brackets. This guidance is especially notable giving research has already differentiated adults by age groups regarding COVID-19 symptoms. Similarly, only adults with small family sizes in regions far from the epicenter were more likely to experience lower levels of life satisfaction, and hence mental healthcare providers can use typhoon eye effect to screen for mental health issues among people with small families.

### Limitations

The context of this study has a clear epicenter of COVID-19, Wuhan, in China. However, not all countries have a single epicenter. For example, South Korea simultaneously had several epicenters ^32^, which opened new research avenues on the effect of geographical distance in the presence of multiple epicenters. In addition, the design was unable to capture the dynamic movement of the citizens across the cities in China over time to observe the fluctuations in the well-being over time. Lastly, our data was collected in China, a geographically large country, and it remains unclear whether the typhoon eye effects will generalize in other countries, most of which are smaller. The epicenter of Wuhan is in the center of the country, yet an epicenter of COVID-19 in the United States of American is New York State, which is geographically on the Northeastern. In that case, we suspect that the typhoon eye effects might play out differently (i.e., at different pace and patterns).

### Conclusion

Our research calls for clinical psychiatrists’ attention on people’s mental health during the pandemic of COVID-19, especially for those who are living away from the epicenter. We found that the further people are away from the epicenter, the lower are their reported life satisfaction. This relationship is stronger among younger individuals and individuals with smaller family sizes but not significant for older adults or those with bigger families. To conclude, our research helps provide insights for clinical professionals in identifying the vulnerable groups of people that are more exposed to the risk of mental disorder during the pandemic of COVID-19.

## Data Availability

Upon request

## Contributors

Stephen X. Zhang contributed to the planning, conduct, and reporting of the work and is responsible for the overall content as the guarantor. Stephen also drafted the empirical sections of the paper and led the submission. Pok Man Tang drafted conceptual parts of the paper. Stephen Zhang, Pok Man Tang, and Chi Hon Li made equal contributions to this paper. Chi Hon Li edited and formatted the manuscript, made the figures and tables, and prepared the submission files. Feng Wei led the data collection and obtained necessary funding and ethical approval. The corresponding author attests that all listed authors meet authorship criteria and that no others meeting the criteria have been omitted.

## Copyright/license for publication

The corresponding author has the right to grant on behalf of all authors and does grant on behalf of all authors, a worldwide license to the Publishers and its licensees in perpetuity, in all forms, formats, and media (whether known now or created in the future), to i) publish, reproduce, distribute, display and store the Contribution, ii) translate the Contribution into other languages, create adaptations, reprints, include within collections and create summaries, extracts and/or, abstracts of the Contribution, iii) create any other derivative work(s) based on the Contribution, iv) to exploit all subsidiary rights in the Contribution, v) the inclusion of electronic links from the Contribution to third party material where-ever it may be located; and, vi) license any third party to do any or all of the above.

## Competing interests

All authors have completed the ICMJE uniform disclosure form at www.icmje.org/coi_disclosure.pdf and declare: no support from any organization for the submitted work; no financial relationships with any organizations that might have an interest in the submitted work in the previous three years; no other relationships or activities that could appear to have influenced the submitted work.

## Transparency declaration

The corresponding author Chi Hon Li affirms that this manuscript is an honest, accurate, and transparent account of the study being reported; that no important aspects of the study have been omitted; and that any discrepancies from the study as planned (and, if relevant, registered) have been explained.

## Funding

The data collection was funded by the Chinese National Funding of Social Sciences (Grant number: 1509093).

## Dissemination declaration

Patients or the public were not involved in the design, or conduct, or reporting, or dissemination plans of our research

## Data sharing

Data are available upon request.

## Notes

### Competing Interest Statement

The authors have declared no competing interest.

### Clinical Trial

Not applicable

## REFERENCES

1. Lagasse J. Healthcare workers risk burnout, exposure in wake of coronavirus pandemic. Healthcare Finance. 2020.

2. Langer G. Coronavirus impacts: Disrupted lives, elevated stress, and soaring worry: Poll. ABC News. 2020.

3. Lombardo P, Jones W, Wang L, et al. The fundamental association between mental health and life satisfaction: results from successive waves of a Canadian national survey. BMC Public Health. 2018; 18: 342.

4. Koivumaa-Honkanen H, Kaprio J, Honkanen RJ, et al. The stability of life satisfaction in a 15-year follow-up of adult Finns healthy at baseline. BMC Psychiatry. 2005; 5: 4.

5. Li S, Rao L-L, Ren X-P, et al. Psychological typhoon eye in the 2008 Wenchuan earthquake. PloS one. 2009; 4:

6. Xie X-F, Stone E, Zheng R, et al. The ‘Typhoon Eye Effect’: determinants of distress during the SARS epidemic. Journal of Risk Research. 2011; 14: 1091–107.

7. Kinsman J. “A time of fear”: local, national, and international responses to a large Ebola outbreak in Uganda. Globalization and Health. 2012; 8: 15.

8. Des Jarlais DC, Galea S, Tracy M, et al. Stigmatization of newly emerging infectious diseases: AIDS and SARS. American Journal of Public Health. 2006; 96: 561–67.

9. Slovic P. Perception of risk. Science. 1987; 236: 280–85.

10. Gao J, Zheng P, Jia Y, et al. Mental health problems and social media exposure during COVID-19 outbreak. Available at SSRN 3541120. 2020:

11. Rafiey H, Momtaz YA, Alipour F, et al. Are older people more vulnerable to long-term impacts of disasters? Clinical Interventions in Aging. 2016; 11: 1791.

12. Lau AL, Chi I, Cummins RA, et al. The SARS (Severe Acute Respiratory Syndrome) pandemic in Hong Kong: Effects on the subjective wellbeing of elderly and younger people. Aging and Mental Health. 2008; 12: 746–60.

13. Greenwood S, Perrin A, Duggan M. Social media update 2016. Pew Research Center. 2016; 11:

14. Phillips DP, Carstensen LL. Clustering of teenage suicides after television news stories about suicide. New England Journal of Medicine. 1986; 315: 685–89.

15. Aptekar L, Boore JA. The emotional effects of disaster on children: A review of the literature. International Journal of Mental Health. 1990; 19: 77–90.

16. Novilla MLB, Barnes MD, Natalie G, et al. Public health perspectives on the family: an ecological approach to promoting health in the family and community. Family & Community Health. 2006; 29: 28–42.

17. Li Q, Guan X, Wu P, et al. Early transmission dynamics in Wuhan, China, of novel coronavirus–infected pneumonia. The New England Journal of Medicine. 2020; 382:

18. World Health Organization. Statement on the second meeting of the International Health Regulations (2005) Emergency Committee regarding the outbreak of novel coronavirus (2019-nCoV), 2020.

19. BBC News. Coronavirus: Wuhan shuts public transport over outbreak. BBC News. 2020.

20. Fifield A, Sun LH. Chinese cities cancel New Year celebrations, travel ban widens in effort to stop coronavirus outbreak. The Washington Post. 2020.

21. Kinetz E. Where did they go? Millions fled Wuhan, China, before coronavirus lockdown. Global News. 2020.

22. Diener E, Emmons RA, Larsen RJ, et al. The satisfaction with life scale. Journal of Personality Assessment. 1985; 49: 71–75.

23. Brody DJ, Pratt LA, Hughes JP. Prevalence of depression among adults aged 20 and over: United States, 2013–2016: US Department of Health and Human Services, Centers for Disease Control and Prevention, National Center for Health Statistics 2018.

24. Halpern-Felsher BL, Millstein SG, Ellen JM, et al. The role of behavioral experience in judging risks. Health Psychology. 2001; 20: 120–29.

25. Kasperson RE, Renn O, Slovic P, et al. The social amplification of risk: A conceptual framework. Risk Analysis. 1988; 8: 177–87.

26. Koivumaa-Honkanen H, Kaprio J, Honkanen R, et al. Life satisfaction and depression in a 15-year follow-up of healthy adults. Social psychiatry and psychiatric epidemiology. 2004; 39: 994–99.

27. Rissanen T, Viinamäki H, Lehto SM, et al. The role of mental health, personality disorders and childhood adversities in relation to life satisfaction in a sample of general population. Nordic Journal of Psychiatry 2013; 67: 109–15.

28. Lariscy RW, Reber BH, Paek H-J. Examination of media channels and types as health information sources for adolescents: Comparisons for black/white, male/female, urban/rural. Journal of Broadcasting & Electronic Media. 2010; 54: 102–20.

29. Sherbourne CD, Hays RD. Marital status, social support, and health transitions in chronic disease patients. Journal of Health and Social Behavior. 1990; 31: 328–43.

30. Lazarus PJ, Jimerson SR, Brock SE. Responding to natural disasters: Helping children and families. National Association of School Psychologists. 2003:

31. Zimmer Z, Kwong J. Family size and support of older adults in urban and rural China: Current effects and future implications. Demography 2003; 40: 23–44.

32. Park C-k. Coronavirus cluster emerges at another South Korean church, as others press ahead with Sunday services. South China Morning Post. 2020.

